# Spatial distribution of erector spinae activity is related to task-specific pain-related fear during a repetitive object lifting task

**DOI:** 10.1101/2021.10.01.21264413

**Authors:** Melanie Liechti, Michael Von Arx, Patric Eichelberger, Christian Bangerter, Michael L. Meier, Stefan Schmid

## Abstract

Fear-avoidance beliefs, particularly the fear of lifting an object with a flexed spine, were shown to be associated with reduced spinal motion during object lifting in both individuals with and without low back pain (LBP). LBP patients thereby also showed potentially clinically relevant changes in the spatial distribution of back muscle activity, but it remains unknown whether such associations are also present in pain-free individuals. The aim of this study was therefore to investigate the relationship between fear-avoidance beliefs and the change in spatial distribution of lumbar paraspinal muscle activity in pain-free individuals during a repetitive object lifting task. Thirty participants completed two pain-related fear questionnaires and performed 25 repetitions of lifting a 5kg-box from a lower to an upper shelf and back, while multi-channel electromyographic signals were recorded bilaterally from the lumbar erector spinae muscles. Changes in spatial distribution were determined by calculating the differences in vertical position of the weighted centroids of muscle activity (centroid shift) between the first and last few repetitions. Multiple linear regression analyses were performed to examine the relationship between the centroid shift and fear-avoidance belief scores. The analyses showed that the fear of lifting an object with a flexed spine was negatively associated with erector spinae activity centroid shift (R^2^ adj. = 0.1832; p = 0.045), which might be an expression of behavioral alterations in order to prevent the back from possible harm.

## 1. INTRODUCTION

Every experience of pain is modulated by emotions, behavior and beliefs [10,30]. Beliefs consist of a concrete conviction of how people think things are, e.g., that pain is a sign of serious injury [5,6,16]. Beliefs underlying pain-related fear are associated with fear of movement or fear of injury, which signifies a fear that a certain movement might be harmful to the body [20,25]. If pain is perceived as threatening, safety behaviors like avoidance of movement might be applied, as described in the fear-avoidance model [26,38].

Back pain related beliefs are often reported in connection with activities involving the lower back such as object lifting [2,7,17,23,28,29]. Lifting with a straight back (spine in neutral position) is thereby commonly believed to be safe, whereas lifting with a round back (flexed spine) is perceived as dangerous [7,29]. However, the biomechanical literature provides no conclusive evidence for justifying the assumption that lifting with a straight back prevents injury and pain [11,12,21,32,37]. On the contrary, keeping the back straight during lifting might lead to muscle fatigue and pain as this technique was shown to result in increased activity of the erector spinae muscles [21].

Results from multi-channel electromyography (EMG) studies reported changes in the spatial distribution of muscle activity over time during different repetitive tasks, which indicated a possible strategy for compensating muscle fatigue in healthy individuals [1,13,14,19]. However, these changes seem to be different in patients with low back pain (LBP), as they demonstrated increased activation of the same muscle regions and significantly less spatial distribution changes than healthy pain-free individuals [1,13]. This reduced muscle activation variability might have important implications regarding provocation and recurrence of LBP [13]. Moreover, research showed a remaining altered trunk muscle recruitment pattern during lifting after remission from a recurrent LBP episode [34], indicating potential long-term modification of muscle recruitment patterns in individuals with a history of LBP.

Interestingly, the higher EMG activity in LBP patients has been shown to be further associated with pain-related fear during a forward bending task, which appeared to be mediated by reduced lumbar flexion [18]. In support of an association between pain-related fear and lumbar flexion, LBP patients showed a significant relationship between reduced lumbar flexion during object lifting and higher task-specific (round back beliefs) but not general pain-related fear [28]. In addition, a recent study revealed that these associations seemed not only to be present in LBP patients but also in healthy pain-free individuals with no history of chronic LBP [22], indicating that pain-related fear might play a role in lifting behavior in the absence of pain.

However, it remains unclear, whether healthy pain-free individuals also show associations between pain-related fear and changes in the spatial distribution of spinal muscle activity during lifting. Such knowledge could substantially contribute to a better understanding of how fear-avoidance beliefs, in particular the fear of lifting an object with a round back, might influence paraspinal muscle activity in a way that could predispose healthy individuals to LBP. For this reason, this study aimed at investigating the relationship between fear-avoidance beliefs and the spatial distribution of erector spinae muscle activity in healthy individuals during a repetitive object lifting task.

## 2. METHODS

### 2.1. Participants

Thirty healthy and pain-free adults (males/females: 20/10; age: 31.8 ± 8.52 years; mass: 71.1 ± 10.2 kg; height: 175.3 ± 7.54 cm; body mass index (BMI): 23.3 ± 2.4 kg/m^2^) were enrolled in this study. Exclusion criteria were any spinal pathologies or surgeries, any musculoskeletal injury that would have limited their function, a BMI ≥ 30, irradiating LBP or LBP affecting activity of daily living during the past six months or if participants were pregnant or in the breastfeeding period. Furthermore, people that were familiar with lifting guidelines such as physical therapists, nurses or anyone practicing CrossFit or Olympic weightlifting were excluded.

The local ethics committee provided exemption for this study (Kantonale Ethikkommission Bern, Req-2020-00364) and all participants provided written informed consent prior to collecting any personal or health related data.

### 2.2. Assessment of pain-related fear

To assess general and task-specific pain-related fear the following two self-report questionnaires were used:

#### 2.2.1. Tampa Scale for Kinesiophobia for the general population (TSK-G)

The TSK-G is a 17-item questionnaire to assess pain-related fear of movement, fear of (re)injury and fear avoidance behavior in the general population without back complaints [20]. The answer scale ranges from “strongly disagree” to “strongly agree”. Four items are phrased in reversed key (items 4, 8, 12, 16) and higher scores on the TSK-G indicate a higher degree of fear of movement. The minimum score lies at 17 points (low kinesiophobia) and the maximal score at 68 points (high kinesiophobia). Psychometric research indicated a sufficient reliability (Cronbach’s α = 0.78) [20]. For this study, we used a modified German version of the TSK-G [22], whereby the total score (TSK-total) served as measure of general pain-related fear.

#### 2.2.1. Photograph Series of Daily Activities-Short electronic Version (PHODA-SeV)

The PHODA-SeV measures the perceived harmfulness of daily activities and movements to the back [25]. It consists of 40 pictures of daily activities on a computer monitor. Participants were requested to imagine themselves performing the shown activity and rate the harmfulness of this activity to their back on a scale ranging from 0 to 100 (0 = “not harmful to the back”, 100 = “extremely harmful to the back”). The reliability and validity of the PHODA-SeV ranged from good to excellent [25]. For the current study, the score of the item three “lifting a pot with a bent back” (PHODA-lift) was used as measure of task-specific pain-related fear. Lifting a pot represents a typical lifting task and previous studies reported a significant relationship of this item with lumbar spine motion in healthy pain-free individuals as well as patients suffering from LBP [22,28].

### 2.3. Biomechanical measurements

#### 2.3.1. Lifting task

To ensure comparability with previous research, we chose the lifting task from Falla et al. [13]. Participants were asked to repetitively lift a box of 5 kg (40 × 23 × 30 cm) between two shelves placed at knee height (lateral epicondyle of the femur with extended knees) and shoulder height (position of the second rib while standing). The box included holes on either side for gripping and a 5kg-dumbbell weight placed in the middle of the box, stabilized by light packing foam. Participants were instructed to lift the box from the lower shelf to the upper shelf in one second, wait for three seconds (still holding the box while it is placed on the shelf) and move the box back to the lower shelf in one second, and wait three seconds before starting the next cycle. In total 25 lifting cycles were performed. A metronome was used to ensure correct timing of the cycles. The high clicking sound of the metronome signaled the start of lifting, whereas the low clicking sound signaled the waiting period. To prevent the electrodes from detaching, participants were asked to avoid excessive forward flexion of their back during the lifting task. For familiarization, participants practiced the lifting task for one minute with the empty box prior to data collection.

#### 2.3.2. Muscle activity

As for the lifting task, the EMG measurement protocol was mainly adapted from the study by Falla et al. [13]. Two multi-channel electrodes (type GR08MM1305; OT Bioelettronica, Torino, Italy) were attached over the left and right lumbar erector spinae muscles. The grid consisted of five columns and thirteen rows of electrodes, with 8 mm interelectrode distance in both directions and with one electrode absent in the upper right corner (Figure 1A).

**Figure 1.**
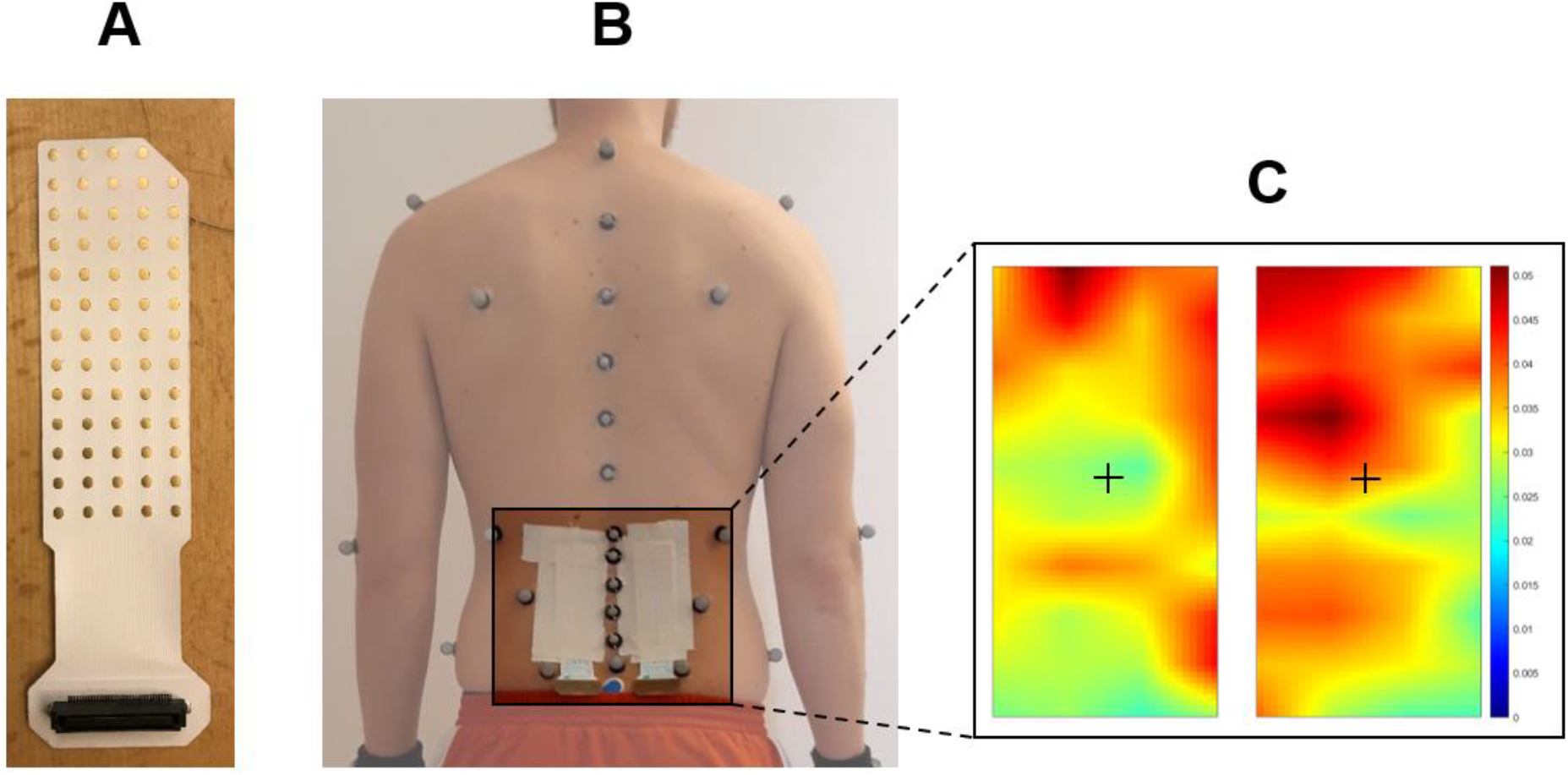
Experimental setup for biomechanical measurements. A) Type of multi-channel electrodes used to derive erector spinae muscle activity. B) Placement of the electrodes over the left and right lumbar erector spinae muscles as well as retro-reflective skin markers over the spinous processes of the lumbar vertebrae and the sacrum. C) Topographical representation of the calculated bipolar erector spinae muscle activity signals (interpolated by a factor 8) as well as weighted centroids (black crosses).

To ensure good skin-electrode contact, the skin was shaved and cleaned with abrasive paste (everi, Spes Medica, Battipaglia, Italy) and alcohol. In addition, a double-sided adhesive foam matrix with 64 holes (Spes Medica, Genova, Italy) was attached to the electrode and the cavities were filled with conductive gel (ac cream, Spes Medica, Genova, Italy). The lower medial corners of the grid were located 2 cm lateral to the fifth lumbar spinous process midpoint bilaterally on the erector spinae muscle (Figure 1B). For the application of the electrodes, participants were asked to stay in a standing position with a slightly flexed lumbar spine. A reference electrode was placed at the sacral bone and a ground electrode at the left lateral malleolus. Monopolar EMG signals were acquired with a sampling frequency of 2048 Hz (EMG-USB2+ Bioelectrical signal amplifier, OT Bioelettronica, Italy, signal gain: 500; built-in bandpass filter: 10-500 Hz).

#### 2.3.3. Kinematics

Participants were equipped with 58 reflective skin markers according to a previously described protocol [33], including markers placed over the spinous processes of L1-L5 as well as on the sacrum on the height of S2 (Figure 1B). Anatomical landmarks were thereby identified by two experienced physiotherapists and markers were attached using double-sided adhesive tape. For orientation and calculation of the start and end point of a lifting cycle, the box was equipped with two additional markers. Three-dimensional marker positions were captured at a rate of 200 frames per second using a 16-camera optical motion capture system (Vicon Motion System Ltd, Oxford, UK).

#### 2.3.4. Data reduction

EMG signals were processed as described in Falla et al. [13] using custom MATLAB routines (version R2020a; Mathworks Inc., Natick, MA, USA). The relevant EMG signal sections were selected based on a one-second window centered around the points of maximum upward and downward velocity (derived from the markers on the box) during the lifting-up and putting-down movements, respectively. To be able to evaluate the change of spatial distribution over the 25 repetitions, signals were extracted from the lifting cycles 2-4 (start phase) and 22-24 (end phase). For each one-second window, 64 monopolar signals for the left and right sides were extracted, bandpass filtered (second order Butterworth; 20-350 Hz) and rectified. Following a comprehensive visual inspection of the quality of the rectified EMG signal for every single channel, we excluded several signals extracted from the top and bottom rows as well as the most lateral columns of the electrodes (i.e. left outer column for the left electrode and right outer column for the right electrode). This resulted in 44 monopolar signals, which were then used to calculate 40 column-oriented differential signals (hereafter referred to as bipolar signals) for each one-second window and side. After calculating the root mean square (RMS) for each bipolar signal over the one-second window, the spatial distribution of EMG activity along the erector spinae muscles was determined by computing the vertical position of a weighted centroid derived from the 40 bipolar signals (Figure 1C). The vertical centroid positions were then averaged over the left and right sides as well as over the start phase and end phase cycles (cycles 2-4 and 22-24, respectively) for the lifting-up and putting-down movements. To investigate potential changes in the spatial distribution over the whole lifting task, the differences in vertical centroid positions between the start and end phases were calculated.

Kinematic data were pre-processed using the software Nexus (version 2.10.3, Vicon Motion System Ltd, Oxford, UK). Pre-processing included the reconstruction and labeling of the markers, the filtering of marker trajectories and the setting of approximate start and end point events for every lifting-up and putting-down movements. For further processing and analysis, a custom-built MATLAB script (version R2020a; Mathworks Inc., Natick, MA, USA) was used. First, an event-detection routine was applied to determine the exact start and end points of the lifting up and putting down movements, which were derived from the displacement of the markers attached to the box. In a second step, we calculated the points of maximum upward and downward velocity of the box, which was used for the analysis of the EMG signals (see above). To determine the lumbar lordosis angles throughout the lifting-up and putting-down movements, we calculated the central angle (in degrees [°]) of a circle fitted into a second-degree polynomial established from the positions of the markers placed over the spinous processes of the lumbar vertebrae and the sacrum as previously described by Schmid et al. [33]. Finally, the continuous lumbar lordosis angles were parameterized as ranges of motion (ROM) and averaged over the start phase and end phase cycles (cycles 2-4 and 22-24, respectively) for the lifting-up and putting-down movements.

### 2.4. Statistical analysis

Statistical analysis was performed using the open-source software jamovi (The jamovi project (2021), version 1.6, www.jamovi.org). Descriptive statistics were calculated for the lumbar lordosis ROM of the start and end phases of the lifting task. Multiple linear regression analyses were performed to examine the relationship between pain-related fear measures and the changes in vertical centroid positions of the erector spinae muscle activity. Normal distribution was confirmed using the Shapiro-Wilk test as well as visual inspection of the Q-Q-plots of the model residuals. In addition, the calculation of Cook’s Distances confirmed the absence of influential cases (outliers). In a first step, a basic model was created containing only the control variables age [3,39,40] and BMI [4,40], since these variables were reported to influence muscle activity as well as its spatial distribution. In a second step, separate regression analyses were performed for every measure of pain-related fear (TSK-total and PHODA-lift) by adding the respective variable to the basic model. This resulted in two additional regression models with the same control variables, but a different measure of pain-related fear for the lifting-up and putting-down phases. Tests corresponded to the null hypothesis which assumed no relationship between pain-related fear measures and the changes in vertical centroid positions of the erector spinae muscle activity. Statistical significance was set at p ≤ 0.05. For statistically significant relationships, effect sizes (f^2^) were calculated based on the adjusted R^2^, with values of ≥ 0.02, ≥ 0.15 and ≥ 0.35 representing small, medium, and large effects, respectively [8].

## 3. RESULTS

### 3.1. Pain-related fear

Mean scores were 32.3 (SD 6.22) for the TSK-total and 42.5 (SD 27.9) for the PHODA-lift. The sum and mean values for all 40 items of the PHODA-SeV are listed in Table 1. The three highest harmfulness ratings of the PHODA-SeV were the a priori selected item PHODA-lift and the items “falling backwards” (PHODA-falling) with a mean of 55.9 (SD 31.6) and “shoveling soil” (PHODA-shoveling) with a mean of 57.7 (SD 28.1). An explorative regression analysis was performed for the latter two items in section 3.3.

**Table 1:**
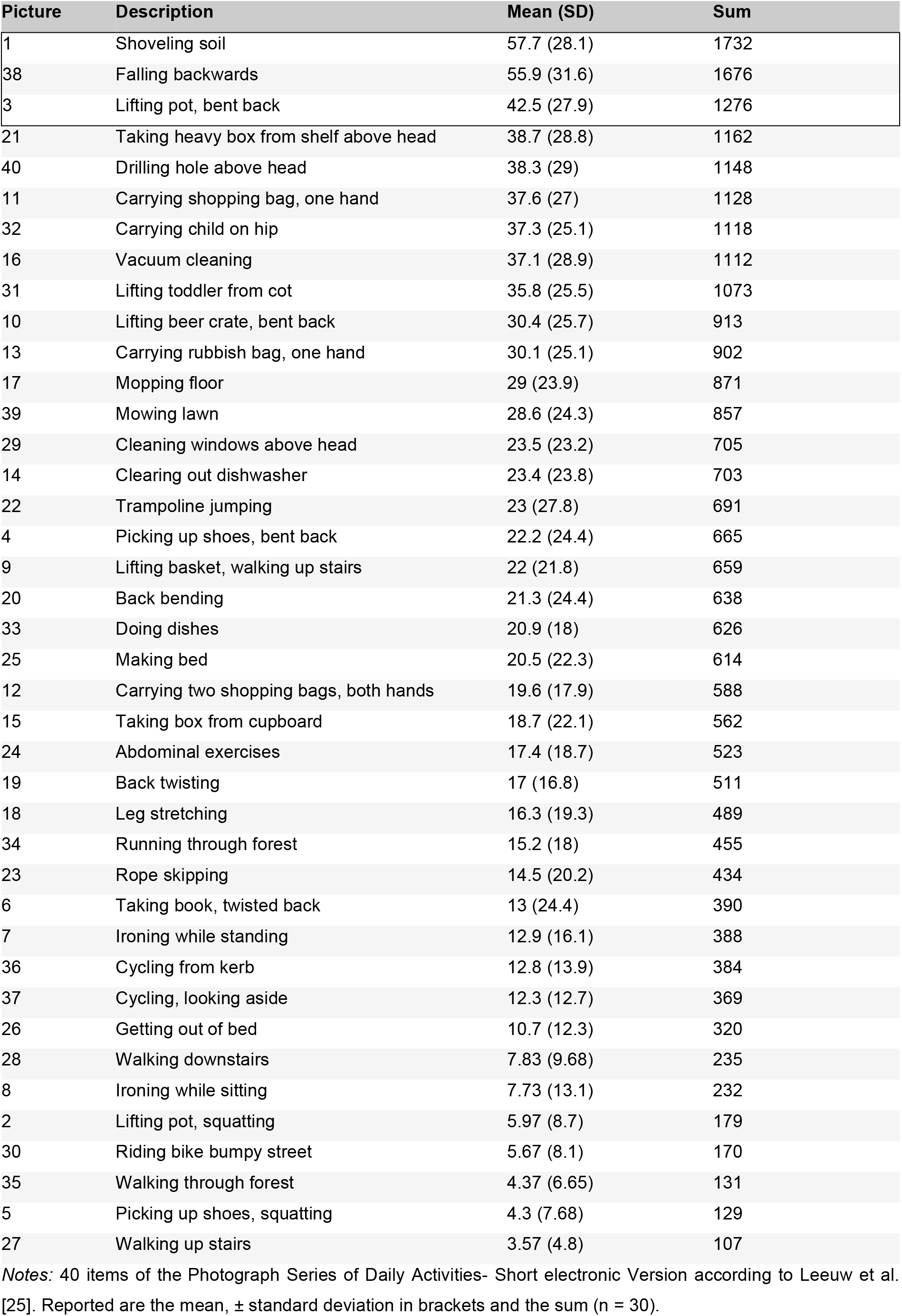
Baseline values of PHODA-SeV organized in descending order of the mean value.

| Picture | Description | Mean (SD) | Sum |
| --- | --- | --- | --- |
| 1 | Shoveling soil | 57.7 (28.1) | 1732 |
| 38 | Falling backwards | 55.9 (31.6) | 1676 |
| 3 | Lifting pot, bent back | 42.5 (27.9) | 1276 |
| 21 | Taking heavy box from shelf above head | 38.7 (28.8) | 1162 |
| 40 | Drilling hole above head | 38.3 (29) | 1148 |
| 11 | Carrying shopping bag, one hand | 37.6 (27) | 1128 |
| 32 | Carrying child on hip | 37.3 (25.1) | 1118 |
| 16 | Vacuum cleaning | 37.1 (28.9) | 1112 |
| 31 | Lifting toddler from cot | 35.8 (25.5) | 1073 |
| 10 | Lifting beer crate, bent back | 30.4 (25.7) | 913 |
| 13 | Carrying rubbish bag, one hand | 30.1 (25.1) | 902 |
| 17 | Mopping floor | 29 (23.9) | 871 |
| 39 | Mowing lawn | 28.6 (24.3) | 857 |
| 29 | Cleaning windows above head | 23.5 (23.2) | 705 |
| 14 | Clearing out dishwasher | 23.4 (23.8) | 703 |
| 22 | Trampoline jumping | 23 (27.8) | 691 |
| 4 | Picking up shoes, bent back | 22.2 (24.4) | 665 |
| 9 | Lifting basket, walking up stairs | 22 (21.8) | 659 |
| 20 | Back bending | 21.3 (24.4) | 638 |
| 33 | Doing dishes | 20.9 (18) | 626 |
| 25 | Making bed | 20.5 (22.3) | 614 |
| 12 | Carrying two shopping bags, both hands | 19.6 (17.9) | 588 |
| 15 | Taking box from cupboard | 18.7 (22.1) | 562 |
| 24 | Abdominal exercises | 17.4 (18.7) | 523 |
| 19 | Back twisting | 17 (16.8) | 511 |
| 18 | Leg stretching | 16.3 (19.3) | 489 |
| 34 | Running through forest | 15.2 (18) | 455 |
| 23 | Rope skipping | 14.5 (20.2) | 434 |
| 6 | Taking book, twisted back | 13 (24.4) | 390 |
| 7 | Ironing while standing | 12.9 (16.1) | 388 |
| 36 | Cycling from kerb | 12.8 (13.9) | 384 |
| 37 | Cycling, looking aside | 12.3 (12.7) | 369 |
| 26 | Getting out of bed | 10.7 (12.3) | 320 |
| 28 | Walking downstairs | 7.83 (9.68) | 235 |
| 8 | Ironing while sitting | 7.73 (13.1) | 232 |
| 2 | Lifting pot, squatting | 5.97 (8.7) | 179 |
| 30 | Riding bike bumpy street | 5.67 (8.1) | 170 |
| 35 | Walking through forest | 4.37 (6.65) | 131 |
| 5 | Picking up shoes, squatting | 4.3 (7.68) | 129 |
| 27 | Walking up stairs | 3.57 (4.8) | 107 |

Notes: 40 items of the Photograph Series of Daily Activities- Short electronic Version according to Leeuw et al. [25]. Reported are the mean, ± standard deviation in brackets and the sum (n = 30).

### 3.2. Relationships between centroid shift and general (TSK-total) as well as task-specific pain-related fear (PHODA-lift)

Results from the regression analyses for the PHODA-lift and TSK-total as variables of interest are presented in Table 2. The BMI in the basic model showed a significant negative relationship with the changes in vertical centroid position of erector spinae muscle activity during the lifting-up phase (stand. estimate = -0.384; p = 0.046). The basic model containing age and BMI showed no relationship with vertical centroid position changes (R^2^ adj. = 0.0861; p = 0.118). Adding the PHODA-lift score to the basic model, a statistically significant negative relationship was observed in the overall model 2 for the lifting-up phase (R^2^ adj. = 0.1832; p = 0.045, f^2^ = 0.22). The PHODA-lift thereby explained an additional 9.7% of variance to the basic model (ΔR^2^ adj. = 0.0971). This negative relationship was also clearly noticeable when looking at the scatterplots (Figure 2A, left). As predictive variable alone, however, the PHODA-lift was not statistically significant (stand. estimate = -0.3596; p = 0.054). Adding TSK-total to the basic model, no statistically significant additional proportion of variance was explained for the lifting-up phase (ΔR^2^ adj. = -0.0365; p = 0.242), which was also reflected by the absence of a consistent trend on the scatterplots (Figure 2B, left). For the putting-down phase, no statistically significant relationships were observed across all variables.

**Table 2:** Regression models displaying the relationships between changes in vertical centroid position of erector spinae muscle activity and general (TSK-total) as well as task-specific pain-related fear (PHODA-lift).

| Movement direction | Regression model | Variables | Stand. Estimate | p-value | R<sup>2</sup> | R<sup>2</sup>adj. | ΔR<sup>2</sup>adj. |
| --- | --- | --- | --- | --- | --- | --- | --- |
| Lifting up | Model 1: Basic model (control variables) |  |  | 0.118 | 0.151 | 0.0861 |  |
|  |  | Age | -0.159 | 0.393 |  |  |  |
|  |  | BMI | -0.384 | 0.046* |  |  |  |
|  | Model 2: Basic model + PHODA-lift |  |  | 0.045* | 0.271 | 0.1832 | 0.0971 |
|  |  | Age | -0.0582 | 0.750 |  |  |  |
|  |  | BMI | -0.3731 | 0.041* |  |  |  |
|  |  | PHODA-lift | -0.3596 | 0.054 |  |  |  |
|  | Model 2: Basic model + TSK-total |  |  | 0.242 | 0.151 | 0.0496 | -0.0365 |
|  |  | Age | -0.15670 | 0.428 |  |  |  |
|  |  | BMI | -0.38397 | 0.051 |  |  |  |
|  |  | TSK-total | 0.00956 | 0.961 |  |  |  |
| Putting down | Model 1: Basic model (control variables) |  |  | 0.354 | 0.0767 | 0.00573 |  |
|  |  | Age | 0.123 | 0.526 |  |  |  |
|  |  | BMI | -0.228 | 0.244 |  |  |  |
|  | Model 2: Basic model + PHODA-lift |  |  | 0.338 | 0.1240 | 0.01883 | 0.0131 |
|  |  | Age | 0.186 | 0.355 |  |  |  |
|  |  | BMI | -0.221 | 0.256 |  |  |  |
|  |  | PHODA-lift | -0.226 | 0.257 |  |  |  |
|  | Model 2: Basic model + TSK-total |  |  | 0.238 | 0.1528 | 0.05118 | 0.04545 |
|  |  | Age | 0.203 | 0.307 |  |  |  |
|  |  | BMI | -0.237 | 0.216 |  |  |  |
|  |  | TSK-total | 0.288 | 0.147 |  |  |  |

The asterisks (\*) indicate statistically significant relationships at the $p \leq 0.05$ level.

**Figure 2.**
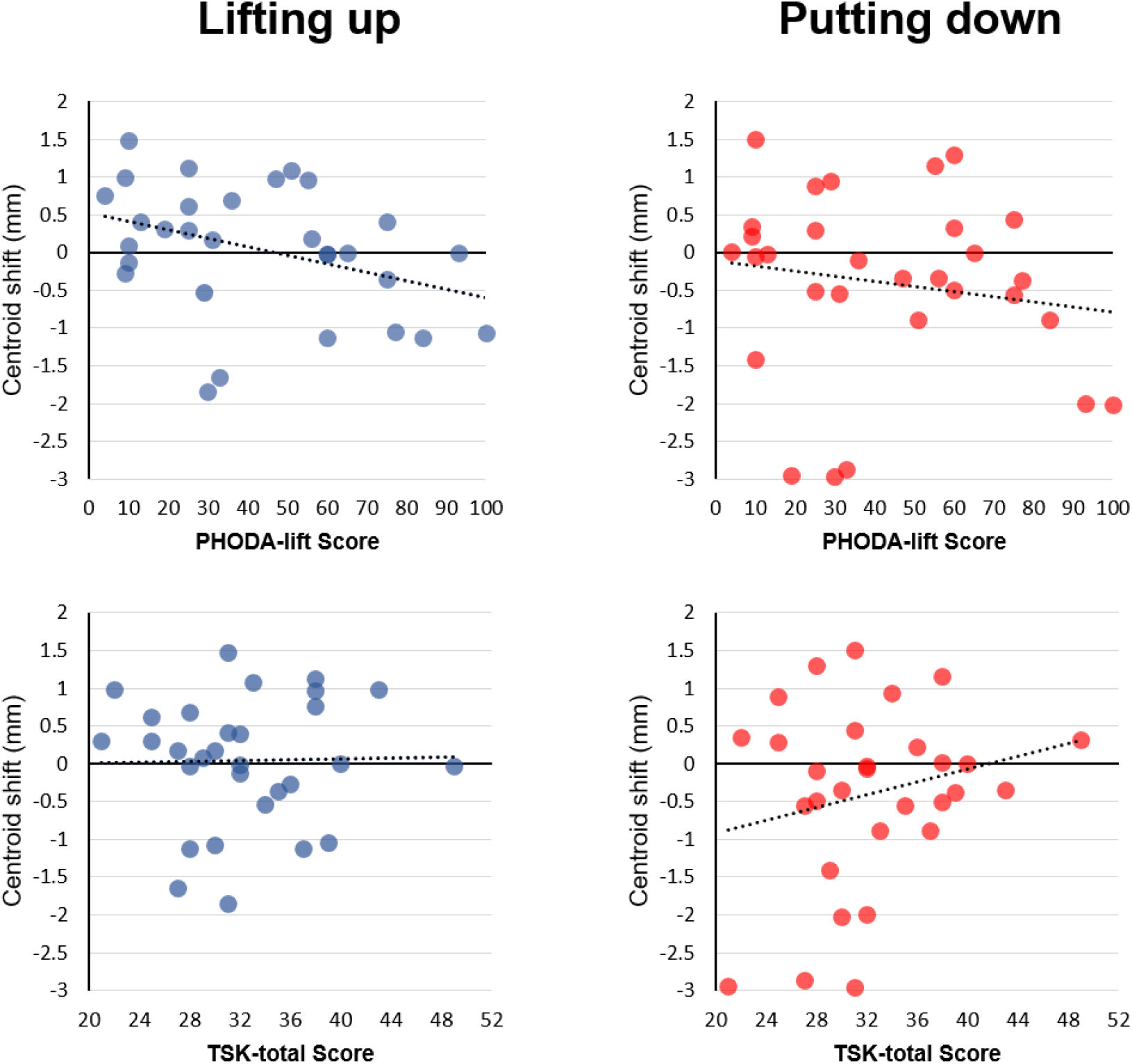
Visualization of the associations between the changes in vertical centroid position of erector spinae muscle activity (centroid shift) and A) task-specific pain-related fear (PHODA-lift) and B) general pain-related fear (TSK-total). The regressions lines are solely displayed for visualization purposes and are not controlled for the variables age and body mass index.

### 3.3. Relationships between centroid shift and additional items of the PHODA-SeV (PHODA-falling and PHODA-shoveling)

Results from the explorative regression analysis for the PHODA-falling and PHODA-shoveling variables are presented in Table 3. The same conditions were applied as in the regression analysis for the PHODA-lift item. In addition to the results from the same basic model, no significant relationship was found between the changes in vertical centroid position of erector spinae muscle activity and the PHODA-falling item. Adding the PHODA-shoveling item to the basic model, a significant relationship for the overall model 2 was observed (R^2^ adj. = 0.1983; p = 0.036, f^2^ = 0.25). PHODA-shoveling explained an additional 11.2% of variance in vertical centroid position changes (ΔR^2^ adj. = 0.1122). Further, PHODA-shoveling, as predictive variable alone, was significantly negatively correlated with the vertical centroid position changes (stand. estimate = -0.367; p = 0.041). No significant relationships were identified between the vertical centroid position changes and the PHODA-falling or PHODA-shoveling items during the putting-down phase.

**Table 3:** Regression models displaying the relationship between changes in vertical centroid position of erector spinae muscle activity and the items PHODA-shoveling as well as PHODA-falling.

| Movement direction | Regression model | Variables | Stand. Estimate | p-value | R<sup>2</sup> | R<sup>2</sup>adj. | ΔR<sup>2</sup>adj. |
| --- | --- | --- | --- | --- | --- | --- | --- |
| Lifting up | Model 1: Basic model (control variables) |  |  | 0.118 | 0.151 | 0.0861 |  |
|  |  | Age | -0.159 | 0.393 |  |  |  |
|  |  | BMI | -0.384 | 0.046* |  |  |  |
|  | Model 2: Basic model + PHODA-falling |  |  | 0.102 | 0.216 | 0.1221 | 0.036 |
|  |  | Age | -0.225 | 0.236 |  |  |  |
|  |  | BMI | -0.434 | 0.026* |  |  |  |
|  |  | PHODA-falling | -0.266 | 0.163 |  |  |  |
|  | Model 2: Basic model + PHODA-shoveling |  |  | 0.036* | 0.284 | 0.1983 | 0.1122 |
|  |  | Age | -0.187 | 0.289 |  |  |  |
|  |  | BMI | -0.425 | 0.021* |  |  |  |
|  |  | PHODA-shoveling | -0.367 | 0.041* |  |  |  |
| Putting down | Model 1: Basic model (control variables) |  |  | 0.354 | 0.0767 | 0.00573 |  |
|  |  | Age | 0.123 | 0.526 |  |  |  |
|  |  | BMI | -0.228 | 0.244 |  |  |  |
|  | Model 2: Basic model + PHODA-falling |  |  | 0.544 | 0.0804 | -0.02991 | -0.0356 |
|  |  | Age | 0.1071 | 0.599 |  |  |  |
|  |  | BMI | -0.2400 | 0.238 |  |  |  |
|  |  | PHODA-falling | -0.0634 | 0.754 |  |  |  |
|  | Model 2: Basic model + PHODA-shoveling |  |  | 0.348 | 0.1213 | 0.01581 | 0.01008 |
|  |  | Age | 0.107 | 0.580 |  |  |  |
|  |  | BMI | -0.252 | 0.201 |  |  |  |
|  |  | PHODA-shoveling | -0.213 | 0.271 |  |  |  |

The asterisks (\*) indicate statistically significant relationships at the $p \leq 0.05$ level.

### 3.4. Lumbar lordosis angle

During the lifting task, participants were able to move their lumbar spine in the sagittal plane. Lumbar lordosis angles during the lifting-up phase showed mean ROMs of 23.8° (SD = 8.05°) and 23.2° (SD = 11.3°) during the start and end phases, respectively. For the putting-down phase, mean ROMs of 21.4° (SD = 6.73°) and 20.9° (SD = 9.51°) were found for the start and end phases, respectively.

## 4. DISCUSSION

The aim of this study was to investigate the relationship between the spatial distribution of erector spinae activity and general as well as task-specific measures of pain-related fear during a repetitive object lifting task. The analysis revealed a significant negative relationship between changes in vertical centroid position of erector spinae muscle activity during the lifting-up phase and task-specific pain-related fear (represented by the PHODA-lift score), whereas general pain-related fear (represented by the TSK-total score) showed no significant relationships.

Results of the TSK-total (mean score 32.3) and PHODA-lift scores (mean score 42.5) indicated similar occurrence of fear avoidance beliefs in healthy pain-free individuals as reported in previous studies [1,7,22]. In addition, comparisons of pain-related fear scores between our study sample involving healthy pain-free individuals and LBP patients in other studies indicated similar scores for TSK-total [1,13,28], but significantly higher scores in LBP patients for PHODA-lift (mean PHODA-lift scores of 77) [28].

Falla et al. [13] investigated the change in distribution of lumbar erector spinae muscle activity by performing a repetitive object lifting task in low back pain and healthy individuals. Results showed a caudal shift of muscle activity distribution in healthy controls over time, whilst the distribution of muscle activity remained unaltered in the LBP individuals. Similar, the results of this study showed a caudal muscle activity centroid shift over time in healthy subjects in presence of high task-specific pain-related fear (PHODA-lift).

In contrast to the PHODA-lift item, the general measure of pain-related fear (TSK-total) showed no significant relationship with the muscle activity centroid shift (R^2^ adj. = 0.0496; p = 0.242). This finding for TSK-total is consistent with some studies [22,28] but not with others [18,31], which is most likely due to a variety of different methodological approaches as well as different population characteristics. In addition, a conditioning process due to task-specific pain experiences might elicit an altered behavioral or muscular reaction. Vlaeyen and Linton [38] showed that an injury could result in increased muscle tension and sympathetic alert, and thus lead to fear. Keeping this in mind, a person might learn to predict potentially harmful events [38], which could lead to a conditioning process resulting in task-specific but not general pain-related fear. However, no data on the occurrence of previous pain events or injuries (exceeding six months prior to the study) were collected in our study.

Furthermore, information obtained or observation of harmful events might trigger a conditioning process [38]. Results from different studies are supportive for this explanatory approach, as beliefs of health care providers were shown to affect their advice given to the patient, with high fear-avoidance beliefs in health care providers being associated with high fear-avoidance beliefs in their patients [9,16]. Information like “lifting with a round back is dangerous” from health care professionals might therefore result in potentially unfavorable beliefs leading to behavioral avoidance [38].

Pointing into this direction, the current findings indicate a relationship between fear avoidance beliefs (in particular the belief that lifting with a round back is dangerous) and the change of muscle activity distribution during a repetitive lifting task, which could be interpreted as a protective muscular response in pain-free individuals.

Results from other multi-channel EMG measures of the erector spinae muscles demonstrated a greater spatial muscle activity distribution with the occurrence of muscle fatigue in both healthy individuals and LBP patients [1,13,35]. However, distribution of muscle activity remained higher in healthy individuals, indicating differences of muscle activity adaption in the presence of LBP [1,13]. In healthy individuals, the larger shifts of muscle activity distribution were associated with longer endurance times and discussed as a strategy to maintain activity [13,14]. However, a study investigated the spatial distribution of erector spinae muscle activity in rowers that recovered from a history of LBP and found reduced distribution of muscle activity and simultaneously a caudal shift of the muscle activity centroid [27]. The caudal shift was attributed to an adapted rowing pattern with altered lumbar flexion, that required higher activation of caudal regions of the erector spinae muscle. In the present study, fear of round-back lifting might induce a protective behavior to avoid lumbar flexion during lifting. To avoid lumbar flexion and remain an upright position of the back, muscular activation of trunk muscles, especially of the erector spinae is required [21,24,27]. This goes along with the results from Knechtle et al. [22], which showed a reduced flexion of the lumbar spine (particularly in the L4/L5 region) in healthy individuals with high task-specific fear during object lifting. This again supports the assumption of movement adaption through a change of muscular activity in the lower lumbar back driven by fear avoidance beliefs.

Besides the PHODA-lift, recent studies showed significant correlations between lumbar spine motion and the item PHODA-falling [22] or PHODA-shoveling [28]. The results of our explorative regression analysis displayed a negative relationship and an additional explanation of variance of 11.2% by the PHODA-shoveling item. This suggests a prediction of the shift of muscle activity towards the caudal direction in presence of high PHODA-shoveling scores with a medium effect size (f^2^ = 0.25). The lifting and shoveling task showed similarities in movement characteristics, as in both tasks the person is handling load with a flexed spine. In contrast, the PHODA-falling item was not related to muscle activity. Falling backwards on the grass (PHODA-falling) might be rated as harmful for the back but does not reflect the same proprioceptive demands as bending the spine and handling loads [28]. In case of conditioning, an external stimulus that resembles the initial painful experience might elicit the same response as to the initial injury [38]. Therefore, PHODA-lift might trigger similar responses in PHODA-shoveling but not in PHODA-falling. Tucker et al. [36] reported that fear avoidance behavior evolved in anticipation of pain and not only in response to experienced pain. They investigated muscle activity pattern in anticipation of pain but without a nociceptive stimulus and reported altered muscle activity recruitment patterns, similar to those observed during periods of pain induced by a direct nociceptive stimulus. They found simultaneous new recruitment and de-recruitment of motor units, and some changes did not resolve as the threat ended [36]. This suggests that the nervous system maintains some adaptations after removal of pain or anticipated pain and therefore might lead to persistent changes in motor control strategies. It was argued that these long-lasting changes in motor unit discharge were not possible due to continuous peripheral nociceptive input, as no direct painful stimulation occurred in the anticipation of pain trials and therefore, central mechanisms must be involved [36]. The peripheral and central mechanisms together could be essential in provocation or recurrence of LBP.

For therapy of LBP, current guidelines recommend education about the nature of LBP and encourage active treatments, which focus on improvements of function and continuation of everyday living activities, addressing peripheral and central aspects of LBP [15]. To improve function, the recognition of task-specific pain-related fear and fear avoidance behavior might be a key factor for therapy and should be further investigated.

There are several limitations of the current study. Spinal motion was restricted, as multi-channel electrodes were attached with rigid tape to the back. Hence, no conclusions about the behavior of lumbar motion were possible. To allow as much unrestricted motion as possible, however, the tape was applied in a supported standing position with a slightly flexed spine. Furthermore, the accuracy of multi-channel electrode signals during a dynamic movement might be compromised by soft tissue elasticity and displacement. In order to minimize noise, the BMI was controlled to exclude individuals with thick fat layers, as previous studies reported this as a significant confounding factor [4,40]. However, measuring the skin fold thickness might provide more specific information about the fat layer on the back as the measurement of BMI does. Another limitation was that the study sample consisted of rather young (mean 31.8 years) and sporty individuals with a mean of 5.4h sports per week, which is not representative for the general population.

In conclusion, results suggest that task-specific pain-related fear contributes to changes in muscle activity distribution of the lumbar erector spinae during lifting. Further investigations are needed to analyze the effect of task-specific pain-related fear in healthy individuals on muscle activity distribution during different tasks of daily living to gain a more comprehensive insight into potential behavioral adaptions as well as future provocation or maintenance of LBP.

## Data Availability

Data will be available upon request.

## 5. ACKNOWLEDGMENTS

The authors thank the volunteers for participating in this study.

## 6. CONFLICT OF INTEREST STATEMENT

The authors have no conflicts of interest to declare.

## Notes

### Competing Interest Statement

The authors have declared no competing interest.

### Funding Statement

No external funding was received for this study.

### Author Declarations

The local ethics committee provided exemption for this study (Kantonale Ethikkommission Bern, Req-2020-00364).

